# Intra-myonuclear inclusions are diagnostic of oculopharyngeal muscular dystrophy

**DOI:** 10.1101/2022.04.25.22274179

**Authors:** Masashi Ogasawara, Nobuyuki Eura, Aritoshi Iida, Theerawat Kumutpongpanich, Narihiro Minami, Ikuya Nonaka, Shinichiro Hayashi, Satoru Noguchi, Ichizo Nishino

## Abstract

The pathologies of oculopharyngeal muscular dystrophy (OPMD) and oculopharyngodistal myopathy (OPDM) are indistinguishable. We found that p62-positive intra-nuclear inclusions (INIs) in myonuclei (myo-INIs) were significantly more frequent in OPMD (11.4 ± 4.1%, range 5.0– 17.5%) than in OPDM and other rimmed vacuolar myopathies (RVMs) (1–2% on average, range 0.0–3.5%, *p*<0.0001). In contrast, INIs in nonmuscle cells (nonmuscle-INIs) were present in OPDM, but absent in other RVMs, including OPMD. These results indicate that OPMD can be differentiated from OPDM and other RVMs by the frequent presence of myo-INIs (≥5%) and the absence of nonmuscle-INIs in muscle pathology.

## INTRODUCTION

Oculopharyngeal muscular dystrophy (OPMD) is a genetic condition that affects muscles and is caused by an alanine expansion mutation in the poly-adenine-binding protein nuclear 1 (*PABPN1*) gene^1^. It is clinically characterized by progressive ptosis, ophthalmoplegia, bulbar muscle involvement, and limb muscle weakness that is predominantly proximal and pathologically by the presence of rimmed vacuoles.^2,3^

Oculopharyngodistal myopathy (OPDM) is a clinicopathologically similar condition to OPMD. It is also characterized by oculopharyngeal muscle involvement and rimmed vacuolar pathology, but the associated limb muscle weakness is typically distal, rather than proximal.^4^ Recently, OPDM was shown to be caused by a CGG repeat expansion in the noncoding region of the *LRP12, GIPC1, NOTCH2NLC*, or *RILPL1* (OPDM_LRP12, OPDM_GIPC1, OPDM_NOTCH2NLC, OPDM_RILPL1) gene.^5-10^

In this study, we characterized the intra-nuclear inclusions (INIs) in muscle specimens from patients with OPMD, three types of OPDM, neuronal intranuclear inclusion disease (NIID), and four other rimmed vacuolar myopathies (RVMs) to pathologically differentiate between OPMD and OPDM—conditions typically indistinguishable by muscle histochemistry.

## METHODS

### Subjects

All samples were sent to the National Center of Neurology and Psychiatry, a referral center for muscle diseases in Japan, for diagnostic purposes. All patients gave informed consent for the use of their samples for research after the diagnosis. We analyzed muscle samples from patients with OPMD (n = 15), OPDM_LRP12 (n = 19), OPDM_GIPC1 (n = 6), OPDM_NOTCH2NLC (n = 7), NIID (n = 10), inclusion body myositis (IBM) (n = 11), GNE myopathy (n = 11), inclusion body myopathy with Paget’s disease of bone and frontotemporal dementia (IBMPFD) with valosin-containing protein gene mutation (n = 8), and DES-related myopathy (n = 7). In addition, we included two control muscle samples without histological abnormalities.

### Muscle histology

Immunohistochemical analysis of anti-p62/SQSTM1 (sc-28359, 1;200, Santa Cruz Biotechnology) was performed using the Ventana immunohistochemistry detection system (Ventana Medical Systems, Tucson, AZ, USA), in addition to hematoxylin and eosin (H&E) staining. We analyzed the frequency of anti-p62 antibody-positive intra-myonuclear inclusions (myo-INIs) in 200 randomly selected myonuclei. Myonuclei strongly stained with anti-p62 were considered to be myo-INIs, but not nuclei with small or >2 dots (Figure 1K),^11^ dots faintly stained (Figure 1L), or those where dots were larger than myonuclei (Figure 1M) to reduce the effects of artifacts and non-specific findings. We further counted the number of p62-positive nuclei in nonmuscle cells (nonmuscle-INIs) such as blood vessels, peripheral nerve bundles, and muscle spindles, except for intrafusal muscle fibers.

**Figure 1.**
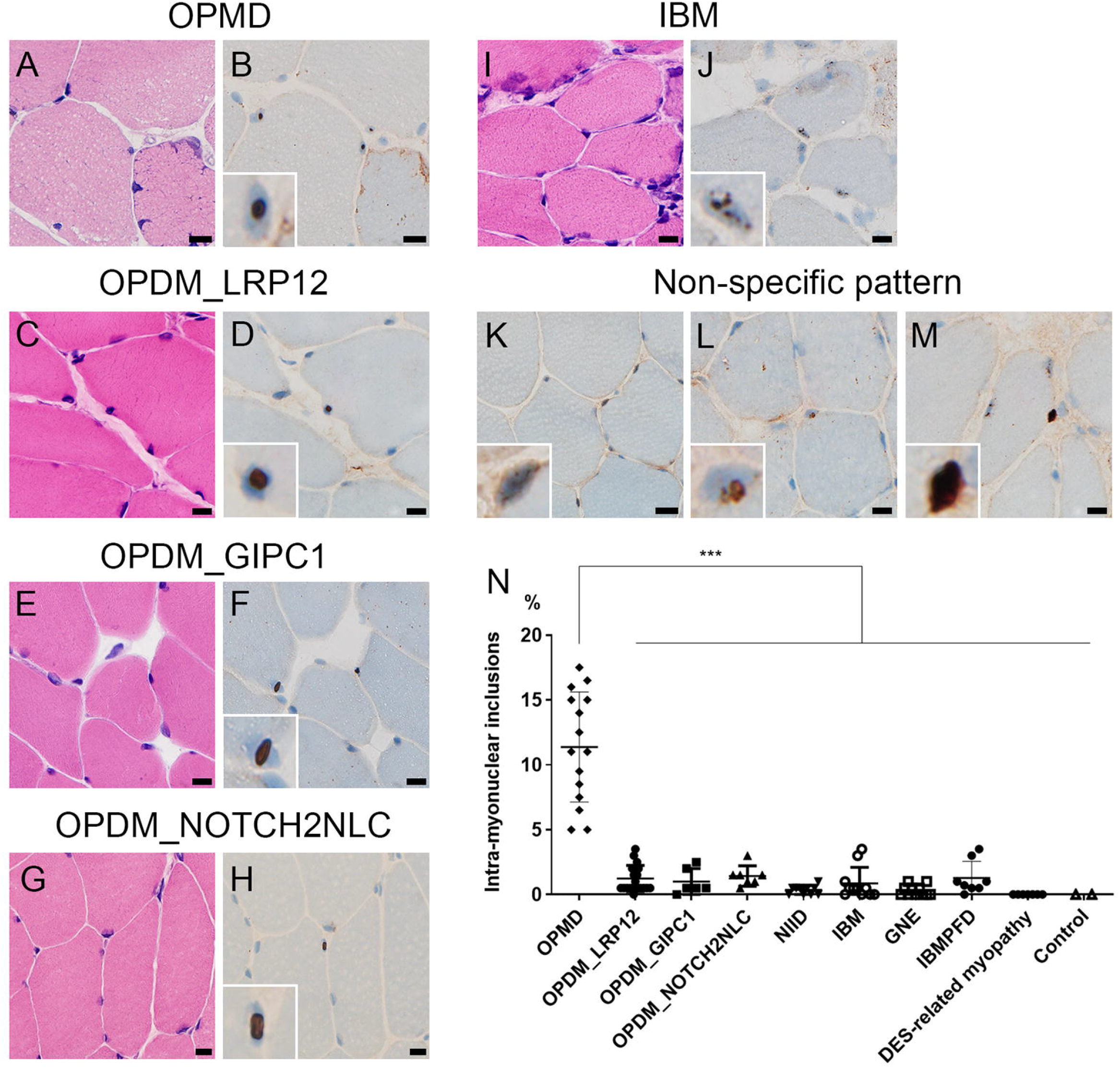
Intra-myonuclei stained by anti-p62 antibody. H&E (A, C, E, G, I) and p-62 antibody (B, D, F, H, J) staining of serial sections of the muscle sample in each disease. Non-specific staining pattern of p-62 antibody in the control (K), GNE myopathy (L), and IBM (M) samples. (N) The graph shows the frequency of p62-positive intra-myonuclei in each disease. (A-M) All black bars show 10 µm. *** OPMD shows higher frequency of p62-positive intra-myonuclei than OPDM_LRP12, OPDM_GIPC1, OPDM_NOTCH2NLC, NIID, IBM, GNE, IBMPFD, DES-related myopathy, and control samples (*p* < 0.0001). H&E: hematoxylin and eosin; IBM: inclusion body myositis; OPMD: oculopharyngeal muscular dystrophy; OPDM: oculopharyngodistal myopathy; NIID: neuronal intranuclear inclusion disease; IBMPFD: inclusion body myopathy with Paget’s disease of bone and frontotemporal dementia

### Electron microscopic analysis

Transmission electron microscopy (EM) was performed by the standard method described previously.^12^

### Statistical analysis

Statistical analyses were performed using GraphPad Prism version 5.03 for Windows (GraphPad Software, San Diego, California, USA). One-way ANOVA with Tukey’s *post hoc* test was used to ascertain the differences of the frequency of p62-positive nuclei among OPMD, OPDM_LRP12, OPDM_GIPC1, OPDM_NOTCH2NLC, NIID, IBM, GNE myopathy, IBMPFD, DES-related myopathy, and the control samples. Results were considered statistically significant at *p* < 0.05.

## RESULTS

In OPMD, the myo-INIs were found in all (100%, 15/15) patients and in 11.4 ± 4.1% (range 5.0 - 17.5%) of myonuclei, while the myo-INIs were absent in the muscle samples taken from the seven DES-related myopathy and the two histologically normal specimens (Figure 1A, B, N, Table 1). In other diseases evaluated in this study, the proportion of myo-INIs varied among patients but were found at a significantly lower frequency than in OPMD: OPDM_LRP12 (1.2 ± 1.0%, range 0.0–3.5%, *p* < 0.0001), OPDM_GIPC1 (1.0 ± 1.0%, range 0.0–2.5%, *p* < 0.0001), OPDM_NOTCH2NLC (1.4 ± 0.7%, range 0.5–3.0%, *p* < 0.0001), NIID (0.4 ± 0.3%, range 0.0– 1.0%, *p* < 0.0001), IBM (0.9 ± 1.2%, range 0.0–3.5%, *p* < 0.0001), GNE myopathy (0.3 ± 0.4%, range 0.0–1.0%, *p* < 0.0001), and IBMPFD (1.3 ± 1.2%, range 0.0–3.5%, *p* < 0.0001) (Figure 1C–J, Table 1). This indicated that OPMD and the other RVMs may be differentiated using a cutoff at 5%. Usually, single INIs were seen in myonuclei in OPMD, three OPDM subtypes, and NIID. In contrast, myonuclei in IBM often contained several smaller p62-positive dots (Figure 1J).

**Table 1.** Anti-p62 antibody-positive intra-nuclear inclusions of muscle fibers and nonmuscle cells in muscle biopsy.

|  | Muscle fiber<br>(% of all myonuclei) | Blood vessel | Schwann cell/<br>pericyte | Perineurium | Spindle |
| --- | --- | --- | --- | --- | --- |
| OPMD | 11.4 ± 4.1% (n=15) | 0 (n=15) | 0 (n=2) | 0 (n=2) | 0 (n=1) |
| OPDM_LRP12 | 1.2 ± 1.0% (n=19) | 0.9 ± 1.4 (n=19) | 1.3 ± 1.4 (n=7) | 1.7 ± 3.3 (n=7) | 3.0 ± 1.4 (n=2) |
| OPDM_GIPC1 | 1.0 ± 1.0% (n=6) | 2.8 ± 3.7 (n=6) | 5.5 ± 2.1 (n=2) | 3.5 ± 0.7 (n=2) | 8.0 ± 1.4 (n=2) |
| OPDM_NOTCH2NLC | 1.4 ± 0.7% (n=7) | 28.1 ± 23.6 (n=7) | 13.6± 13.3 (n=5) | 13.0 ± 11.7 (n=5) | 5 ± 1.4 (n=2) |
| NIID | 0.4 ± 0.3% (n=10) | 26.0 ± 33.1 (n=10) | 9 ± 14.7 (n=3) | 5.7 ± 4.7(n=3) | 1 (n=1) |
| IBM | 0.9 ± 1.2% (n=11) | 0.2 ± 0.6 (n=11) | 0.3 ± 0.5(n=4) | 0 (n=4) | 0 (n=3) |
| GNE myopathy | 0.3 ± 0.4% (n=11) | 0 (n=10) | 0.3 ± 0.8 (n=6) | 0.5 ± 0.7 (n=6) | 0 (n=2) |
| IBMPFD | 1.3 ± 1.2% (n=8) | 0.1 (n=8) | 0 (n=1) | 1 (n=1) | 0 (n=1) |
| DES-related myopathy | 0.0 ± 0.0% (n=7) | 0 (n=7) | 0 (n=3) | 0 (n=3) | NA |
| Control | 0.0 ± 0.0% (n=2) | 0 (n=2) | NA | NA | NA |

OPMD: oculopharyngeal muscular dystrophy; OPDM: oculopharyngodistal myopathy; NIID: neuronal intranuclear inclusion disease;
IBMPFD: inclusion body myopathy with Paget's disease of bone and frontotemporal dementia; IBM: inclusion body myositis

We further evaluated non-muscle-INIs distributed in the blood vessels, Schwann cells/pericytes, perineurium, and muscle spindles, except the intrafusal muscle fibers in muscle biopsy samples. None of the samples from patients with OPMD (n = 15) or DES-related myopathy (n = 7) or the control muscle samples (n = 2) showed nonmuscle-INIs (Table 1). In contrast, nonmuscle-INIs were observed in 53% (10/19), 67% (4/6), 100% (7/7), 100% (10/10), 18% (2/11), 10% (1/10), and 25% (2/8) of patients with OPDM_LRP12, OPDM_GIPC1, OPDM_NOTCH2NLC, NIID, IBM, GNE myopathy, and IBMPFD, respectively (Figure 2A-Y). The number of nonmuscle-INIs was significantly higher in OPDM_NOTCH2NLC (48.6 ± 41.5 [n=7]) than in OPDM_LRP12 (2.4 ± 3.3 [n=19], p <0.0001), OPDM_GIPC1 (8.5 ± 11.3 [n=6], p =0.0034), IBM (0.27 ± 0.6 [n=11], p <0.0001), GNE myopathy (0.3 ± 0.9 [n=10], p <0.0001), and IBMPFD (0.3 ± 0.4 [n=8], p <0.0001) (Figure 2Y). Of note, the number of nonmuscle-INIs found in two IBM, one GNE myopathy, and two IBMPFD patients was ≤ 3 (Table 1). There was no significant difference in the number of INIs among patients with OPDM_GIPC1 and OPDM_LRP12, IBM, GNE myopathy, IBMPFD, and DES-related myopathy and the control samples.

**Figure 2.**
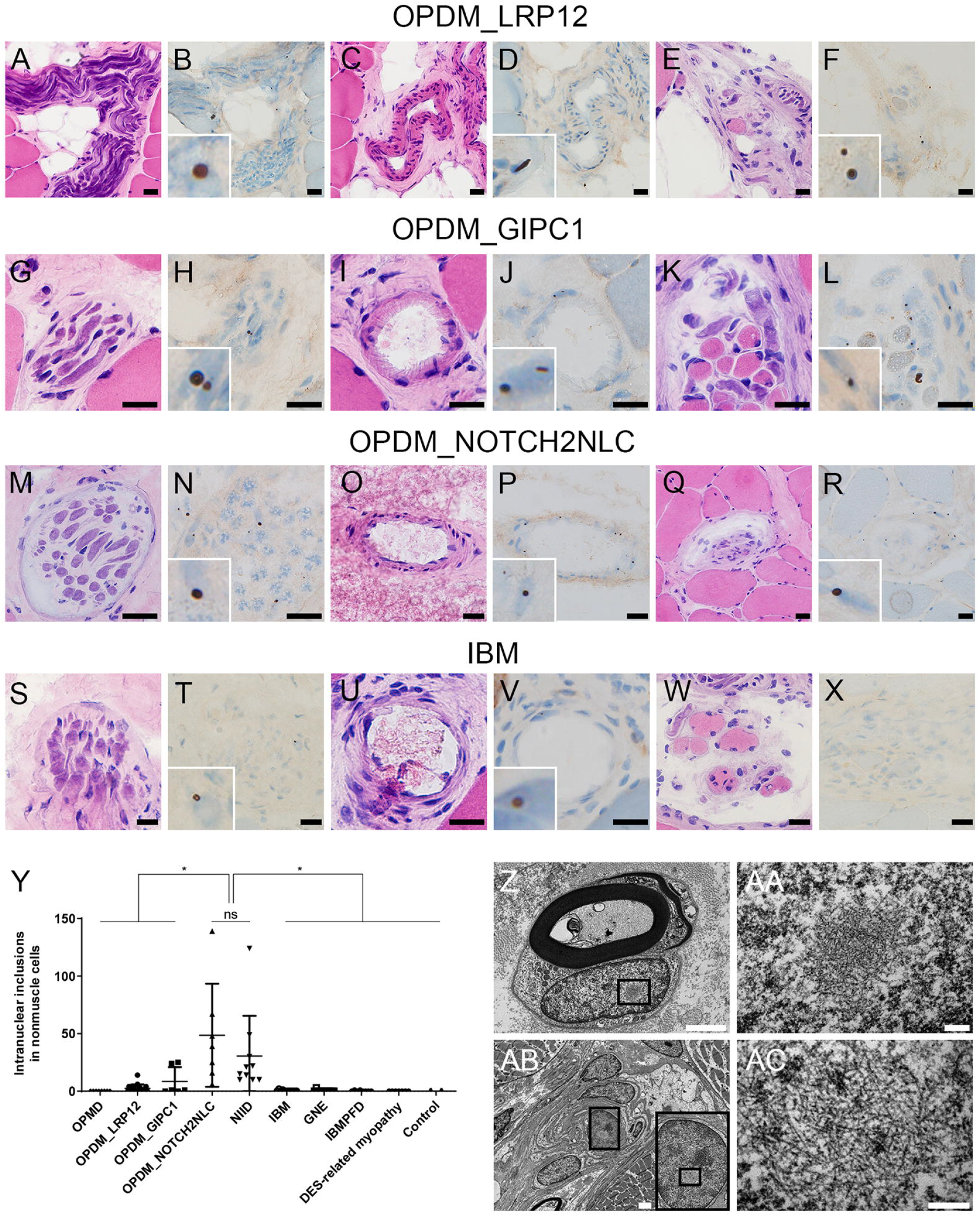
Intra-nuclear inclusions of nonmuscle cells in muscle biopsy. H&E (A, C, E, G, I, K, M, O, Q, S, U, W) and p62 (B, D, F, H, J, L, N, P, R, T, V, X) staining of serial sections of the muscle sample in each disease. The p62-positive INIs are observed in Schwann cells, pericytes, and/or perineurium cells (B, H, N, T), blood vessels (D, J, P, V), and muscle spindles (F, L, R). The p62-positive INIs are not seen in muscle spindles in IBM (W, X). (A-X) All black bars show 20 µm. (Y) Frequency of p62-positive INIs in nonmuscle cells in total in each disease. (Z-AC) With EM, tubulofilamentous inclusions within the Schwann cells (Z, AA) and pericytes (AB, AC) are observed in OPDM_GIPC1. (Z, AB) White bars show 2 µm. (AA, AC) White bars show 200 nm. (Z, AB) Black boxes are magnified. *, p < 0.05; **, p < 0.01; ***, p < 0.0001; ns, no significance. H&E: hematoxylin and eosin; INIs: intra-nuclear inclusions; IBM: inclusion body myositis; EM: electron microscopy; OPDM: oculopharyngodistal myopathy; NIID: neuronal intranuclear inclusion disease; IBMPFD: inclusion body myopathy with Paget’s disease of bone and frontotemporal dementia; OPMD: oculopharyngeal muscular dystrophy

Using EM, tubulofilamentous inclusions (TFIs), without limiting membrane but with low-electron density halo around the nucleus, were observed in the Schwann cells and pericytes of the muscle from a patient with OPDM_GIPC1 (Figure 2Z-AC).

## DISCUSSION

In muscle histochemistry, OPMD and OPDM show indistinguishable features, characterized by the presence of rimmed vacuoles in addition to angular-shaped atrophic fibers.^3, 4, 13^ Furthermore, both conditions present with adult-onset progressive ptosis, ophthalmoplegia, bulbar symptoms, and limb muscle weakness, although proximal limb muscles are predominantly affected in OPMD and distal limb muscles in OPDM. Nevertheless, some of the OPMD patients are known to develop OPDM phenotype, while some OPDM patients present predominantly proximal muscle involvement, making the diagnosis even more difficult without genetic information.^7, 13-15^ In this study, the frequency of myo-INIs was found to be higher in OPMD (11.4 ± 4.1%, range 5.0–17.5%) than in the three types of OPDM (1-2% on average, range 0–3.5%, *p* < 0.0001), suggesting that these conditions could be differentiated by setting a cutoff at the frequency of 5%. No nonmuscle-INIs were seen in OPMD samples, but they were seen in all patients with OPDM_NOTCH2NLC and NIID, and some with OPDM_GIPC1 and OPDM_LRP12. These results suggest that the high frequency (≥5%) of myo-INIs together with the absence of nonmuscle-INIs could differentiate OPMD from OPDM in muscle pathology.

We have recently reported that INIs in skin biopsy samples are found not only in NIID, but also in three types of OPDMs, suggesting that the underlying pathogenesis of OPDM could be identical to that of NIID, as these diseases are caused by a CGG repeat expansion.^16^ In contrast, here, the frequency of the nonmuscle-INIs in muscle biopsy samples was significantly higher in OPDM_NOTCH2NLC than in OPDM_GIPC1 and OPDM_LRP12. However, the frequency of myo-INIs was similar among the subtypes of OPDM, indicating that the CGG expansions in the *NOTCH2NLC* gene affect a wider range of cell types, which may help differentiate OPDM_NOCTH2NLC from other OPDM subtypes in muscle pathology.

Although patients with IBM, GNE myopathy, and IBMPFD also showed nonmuscle-INIs, the number of positive nuclei were ≤ 3. This indicated that the development of nonmuscle-INIs is unlikely to be a mainstream pathologic process in these conditions.

In conclusion, OPMD can be pathologically differentiated from OPDM and other RVMs by the frequent detection of myo-INIs (≥5%) and the absence of nonmuscle-INIs.

## Data Availability

All data produced in the present study are available upon reasonable request to the authors

## Data availability

The data supporting the findings in this study are available from the corresponding author upon request.

## Ethics statement

All patients provided informed consent for using their samples for research after the diagnosis. This study was approved by the ethical committees of the National Center of Neurology and Psychiatry (A2019-123).

## Acknowledgments

We would like to thank all attending physicians and patients for sending muscle and blood samples to us. This study was supported partly by the Intramural Research Grant (2-5) for Neurological and Psychiatric Disorders of NCNP and AMED under Grant Number JP21ek0109490h0002.

## Author contributions

MO and IN designed the studies; AI and NM performed genetic diagnosis; MO, NE, TK, IN, SH, SN, and IN evaluated muscle pathology; MO, NE, AI, SH, SN, and IN wrote and edited the manuscript; and IN supervised the project. All authors read and approved the final manuscript.

